# Evaluation of vaccine-induced binding and neutralizing antibodies as correlates of risk of HPV

**DOI:** 10.1101/2025.05.28.25328434

**Authors:** Zoe Moodie, Bhavesh Borate, Youyi Fong, Partha Basu, Richard Muwonge

**Author notes:** Corresponding author: Zoe Moodie.

## Abstract

**Background:** Estimated vaccine efficacy (VE) at 15 years for a single dose of quadrivalent human papillomavirus (HPV) vaccine in adolescent girls in India exceeded 90% against persistent infection (PI) from HPV 6/11/16/18, similar to VE for two or three doses [1]. Binding and neutralizing antibodies were evaluated as correlates of risk.

**Methods:** HPV-specific antibody titers were measured by ELISA and pseudovirion-based neutralization assay on 25 PI cases of HPV 6, 11, 16, 18 and/or 31 and on 126 matched controls. HPV 31 was included, given cross-protection, to provide sufficient cases for analysis; limited numbers prevented assessment of correlates to each type. Age-adjusted logistic regression models assessed antibody biomarkers as correlates.

**Results:** HPV 18 binding antibody titers inversely correlated with PI: OR=0.62, 95%CI=(0.39, 0.99), p=0.045 per 10-fold titer increase. Our analysis identified no other correlate.

**Conclusion:** Higher HPV 18 titers were associated with a lower risk of PI with HPV 6/11/16/18/31.

The clinical trial is registered with ISRCTN, ISRCTN98283094, and ClinicalTrials.gov, NCT00923702.

## Introduction

Single-dose HPV vaccines were recommended by the WHO in 2022 to expand global access to prevent an estimated 60 million cervical cancer cases and 45 million deaths by 2122 [2]. As of February 4, 2025, 67 countries include a single dose HPV vaccination programme [3]. The WHO recommendation was based in part on the high estimated VE against persistent HPV 16/18 infection over 18 months of follow-up in the KEN-SHE trial: 97.5% (95%CI 81.7 or 81.6% to 99.7%) for single dose of bivalent or nonavalent HPV vaccine [4]. In addition, 9-14 year olds in Tanzania had non-inferior HPV 16 and 18 IgG binding antibody responses 2 years post-first dose to those seen in the Costa Rica Vaccine trial and International Agency for Research on Cancer [IARC] India trial showing high VE of the single-dose regimen [5].

While antibodies are thought to be primary mediators of protection, they have not been established as a correlate of protection in the HPV setting, which would aid in bridging to other populations/vaccine formulations, shortening trial duration, and providing confidence around durability of protection. A better understanding of the implications of lower levels of antibody titers is also needed as protection has been seen even among girls with low neutralizing antibody titers [6–9]. Compared to multi-dose regimens, single dose regimens have shown lower but durable levels of antibodies. Given the high VE, it can take decades to accrue sufficient breakthrough HPV infections to ensure sufficient power for a correlates assessment. The first opportunity for this assessment is provided by the IARC India trial of 1, 2, and 3 doses of quadrivalent HPV vaccine (Gardasil [Merck Sharp & Dohme, Whitehouse Station, NJ, USA]) where post-vaccination blood samples were collected on all participants.

We evaluated HPV-specific binding and neutralizing antibodies measured 18 months post-first vaccination as correlates of risk of persistent HPV infection in the IARC India trial.

## Methods

### Trial design

We conducted a case-control analysis to assess correlates of risk using data from a multicenter, cohort study to evaluate vaccine efficacy of the quadrivalent HPV vaccine among unmarried girls aged 10–18 years in India initiated on September 1, 2009. The trial methods and results have been published [10]. Briefly, estimated vaccine efficacy (VE) at median 12 years of follow-up against persistent HPV 6, 11, 16 and 18 infections was 87.0% (95% CI 81.2% to 91.0%) for 1 dose, 88.0% (95% CI 81.6% to 92.2%) for 2 doses, and 86.9% (95% CI 80.6% to 91.2%) for 3 doses. [1]. Blood samples were collected 18 months after enrollment in all participants.

HPV genotype assessment was performed once participants entered the cervical sampling cohort: 18 months after marriage or 6 months after the birth of their first child, whichever event occurred first. Cervical samples were collected at cohort entry then annually for ≥ 3 samples. If a new infection was detected at the last planned cervical sample collection, then a sample was collected the following year to assess persistent infection. Persistent infection is defined as the detection of same HPV genotype by the Luminex assay (Luminex, Austin, TX, USA) in two consecutive yearly cervical samples with no intervening negative test.

### Study participants

The main analysis population consists of all participants who: a). received at least one dose of vaccine, b). were eligible and enrolled in the cervical sampling cohort after Month 18, c). were followed at least 3 years post-entry into the cervical sampling cohort with annual cervical sampling, and d). were either cases or controls. Cases were participants in the main analysis population with persistent infection with HPV 6, 11, 16, 18, and/or 31 after Month 18. We refer to these as “Any HPV” cases. Controls were participants in the main analysis population who were never detected with incident infection of any of the 21 HPV genotypes assessed: HPV 6, 11, 16, 18, 26, 31, 33, 35, 39, 45, 51, 52, 53, 56, 58, 59, 66, 68, 70, 73, 82.

An alternative analysis population was also considered where only participants who were susceptible (defined as married or pregnant) after Month 18 were included, rather than including all eligible and enrolled participants as in condition b). above. This excluded participants who were married or pregnant (i.e., potentially exposed) prior to the Month 18 immune response assessment. Date of pregnancy was calculated as delivery date minus 9 months.

### Outcome variables

Month 18 antibody responses were measured in all cases detected by December 21, 2019 and in controls with Month 18 samples available. For each case, 5 controls were randomly selected with frequency-matching based on age at enrollment (≤14, > 14), dose group (1 [at month 0 only], 2 default [at 0, 2 months], 2 planned [at 0, 6 months], 3 [at 0, 2, 6 months]), and site (Mizoram/non-Mizoram, where Mizoram had higher HPV incidence).

Total IgG binding antibody titers to HPV types 6, 11, 16, 18, 31, 33, 45, 52, 58 were measured by the multiplex HPV virus-like particle-based IgG ELISA on Meso Scale Discovery platform (M9ELISA) at Centers for Disease Control and Prevention (CDC, Atlanta, USA) laboratory on all HPV 6, 11, 16, 18 and/or 31 (“any HPV”) persistent infections and their matched controls [11].

Neutralizing antibody titers against HPV-L1 protein for HPV types 6, 11, 16, 18, 31, 33, 45, 52, 58 were measured by high throughput pseudovirion-based neutralizing assay (HT-PBNA) at the German Cancer Research Center (DKFZ) Heidelberg, Germany on the same case-control cohort [11].

### Statistical Methods

Logistic regression models of persistent infection with “Any HPV” were fit for each biomarker, adjusting for age>14, using the maximum likelihood estimation method for case-control samples from the ‘osDesign’ package in R statistical software (version 4.3.3; R Foundation for Statistical Computing, Vienna, Austria). Inverse sampling probability weights were computed by stratifying on treatment and age group to account for the case-control sampling.

Each biomarker was studied as a continuous variable and as a categorical variable based on tertiles (low/medium/high) or LLOQ (low/high) for response rates < 1/3.

Multiple testing adjustment via q-values for the false discovery rate and by Holm method for the family-wise error rate was performed across the two primary biomarker outcomes: continuous and categorical scores. Scores were defined by combining the 5 biomarkers by maximum signal diversity weight [12], aiming to provide a parsimonious representation of multiple latent signals. This approach was designed to enhance power in low-sample-size settings for detecting correlations between clinical outcomes and biomarkers.

Two-sided p-values are reported with p-values<0.05 deemed statistically significant.

R statistical software (version 4.3.3; R Foundation for Statistical Computing, Vienna, Austria) was used for statistical analysis.

### Ethical considerations

Every participant provided a written informed consent. The study was approved by the site ethics committees and ethics committee at IARC. The trial is registered with ClinicalTrials.gov (NCT00923702).

## Results

### Baseline characteristics

Of the 4010 participants in the main analysis population, the average age at first vaccination was 15 years with range from 10 to 18 years, 2371 (59.1%) were over the age of 14 years, and with enrollment occurring at 9 sites in India (Table S1).

### Persistent HPV infections

All cases had persistent infection detected at the first assessment, time of entry into the cervical sampling cohort. HPV 31 was the most commonly detected persistent infection (18/35=51.4%), followed by HPV 18 (9/35=25.7%), HPV 6 (4/35=11.4%), HPV 11 and 16 (each 2/35=5.7%).

Month 18 binding and neutralizing antibody titers were available on 25/35 and 26/35 cases respectively and on 126/3952 and 128/3952 controls respectively in the primary analysis population. In the alternative analysis population, these are available on 22/26 cases (both for binding and neutralizing antibodies) and on 109/3175 and 110/3175 controls respectively. The numbers of type-specific cases with binding and neutralizing antibody data in the main analysis population are shown in Figure 1.

**Figure 1:**
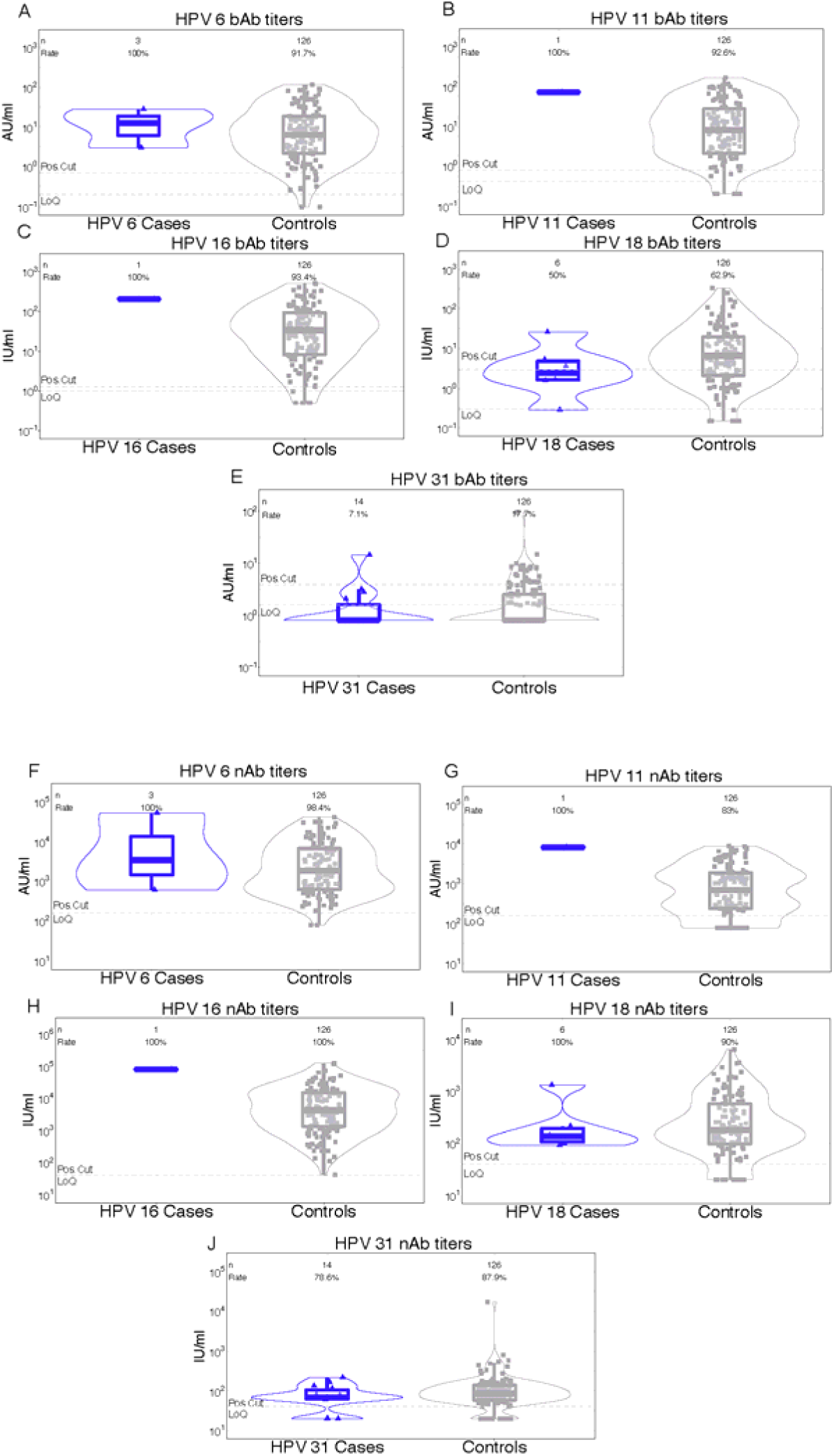
Distribution of Month 18 binding (A-E) and neutralizing (F-J) antibody titers to HPV 6, 11, 16, 18, 31 for type-matched HPV cases and controls in the main analysis population. The number of participants is denoted by n; Rate refers to the proportion of participants with a positive response; Pos. Cut denotes the positivity cutoff; LoQ denotes the limit of quantitation.

### Month 18 binding and neutralizing antibody responses

High correlations (0.74-0.92) were observed between binding antibody responses to each possible pair of quadrivalent vaccine-targeted HPV types, with moderate correlation (0.54-0.57) between HPV 31 and the vaccine-targeted types (Figure S2). Similar results were seen for neutralizing antibody responses (Figure S3).

Binding antibody titer distributions were similar between cases (both Any HPV and type-specific) and controls for the vaccine-targeted HPV types and other types assessed, apart from HPV 18 titers (Figures 1A-E, 2, S4). HPV 18 titers were slightly lower among Any HPV cases with a seropositive rate of 48% compared to 63% among controls (Figure 2D) and also lower among HPV 18 cases with a seropositive rate of 50% compared to 63% among controls (Figure 1D). Neutralizing antibody distributions were also similar between cases (both Any HPV and type-specific) and controls (Figures 1, S1, S5).

**Figure 2:**
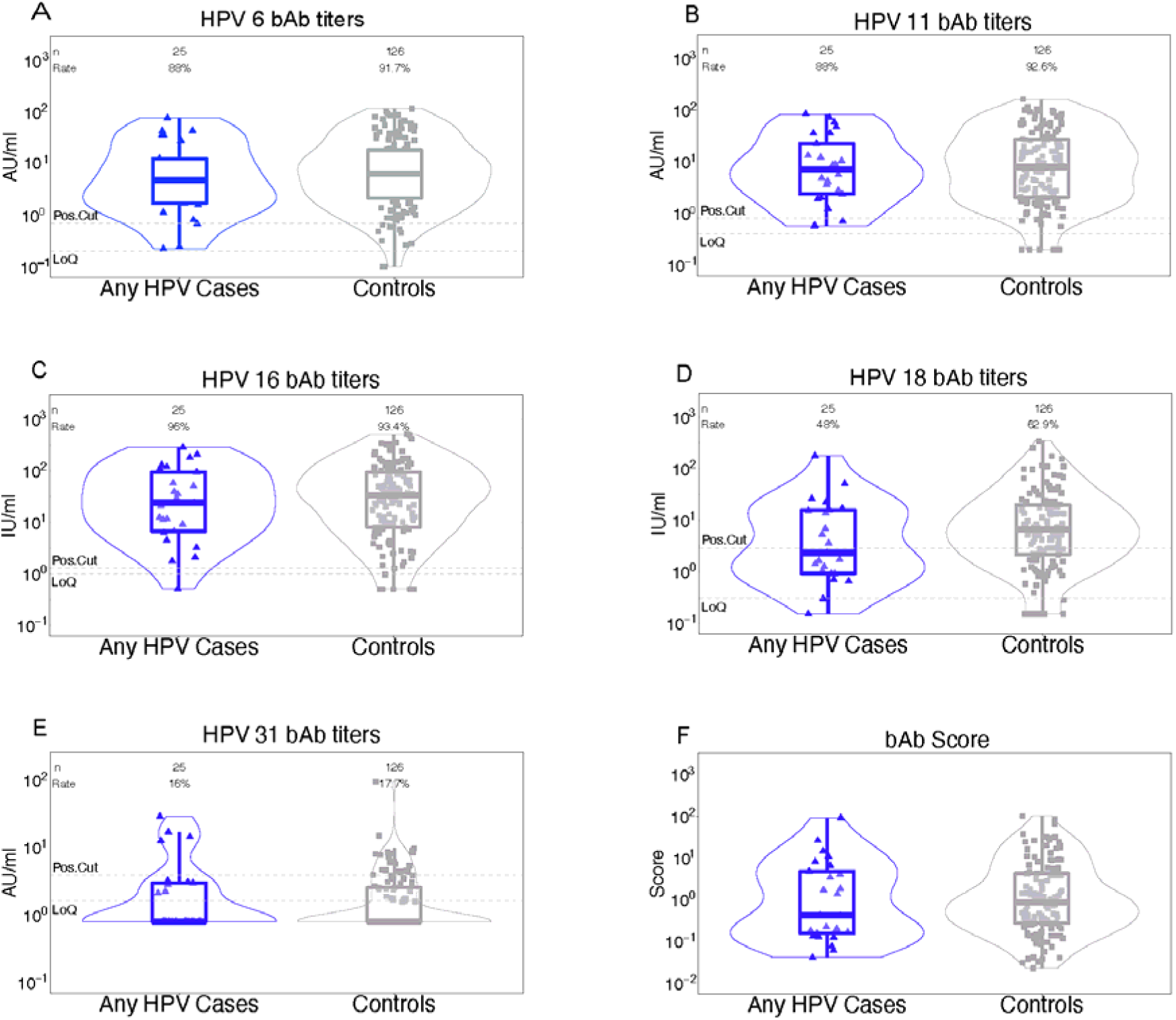
Distribution of Month 18 binding antibody titers to HPV 6, 11, 16, 18, 31 and the score for Any HPV cases and controls in the main analysis population. The number of participants is denoted by n; Rate refers to the proportion of participants with a positive response; Pos. Cut denotes the positivity cutoff; LoQ denotes the limit of quantitation.

There was evidence to support HPV 18 binding antibody titer as an inverse correlate of risk for both continuous and categorical marker levels in the main analysis population (eligible and enrolled in the cervical sampling cohort after month 18). Higher HPV 18 binding antibody titers were associated with a lower odds of Any HPV: OR=0.62, 95%CI=(0.39, 0.99), p=0.045 for a 10-fold increase in HPV 18 titer (Table 1), OR=0.21, 95%CI=(0.06, 0.74), p=0.02 for medium vs. low levels of HPV 18 and OR=0.57, 95%CI=(0.26, 1.23), p=0.15 for high vs. low levels of HPV 18 (Table 2). Similar results were also seen in the alternative analysis population of participants susceptible after Month 18 (Tables S1-S2). Although the number of HPV 18 cases with month 18 titers (n=6) was too small for formal correlates assessment, Figure 1D suggests an association may exist: the 75^th^ percentile of HPV 18 binding antibodies among HPV 18 cases was less than the median among controls.

**Table 1:** Continuous Month 18 IgG binding antibody markers as correlates of risk of Any HPV in the main analysis population (eligible and enrolled in cervical sampling cohort after Month 18). Odds ratios (ORs) are reported from univariate logistic regression models of the marker, adjusted for age > 14 and accounting for case-control sampling via inverse sampling probability weights. Shaded row indicates p-value<0.05 (p-value=0.045 prior to rounding).

| Marker | # cases/# at risk | OR per 10-fold increment (95% CI) | p-value |
| --- | --- | --- | --- |
| Score<sup>1</sup> | 35/3987 | 0.89 (0.54, 1.45) | 0.64<sup>2</sup> |
| HPV 6 | 35/3987 | 0.75 (0.43, 1.29) | 0.30 |
| HPV 11 | 35/3987 | 0.89 (0.52, 1.50) | 0.65 |
| HPV 16 | 35/3987 | 0.85 (0.49, 1.45) | 0.54 |
| HPV 18 | 35/3987 | 0.62 (0.39, 0.99) | 0.05 |
| HPV 31 | 35/3987 | 1.45 (0.53, 3.97) | 0.47 |
| HPV 33 | 35/3987 | 3.21 (0.01, >100) | 0.71 |
| HPV 45 | 35/3987 | 0.20 (0.03, 2.33) | 0.23 |
| HPV 52 | 35/3987 | 1.25 (0.25, 6.26) | 0.78 |
| HPV 58 | 35/3987 | 1.23 (0.22, 6.89) | 0.81 |

<sup>1</sup>The score combined the 5 primary biomarkers (HPV 6, 11, 16, 18, 31) by maximum signal diversity weight, aiming to provide a parsimonious representation of multiple latent signals.
<sup>2</sup>Overall q-value=0.64, overall FWER-adjusted p-value=0.64

**Table 2:**
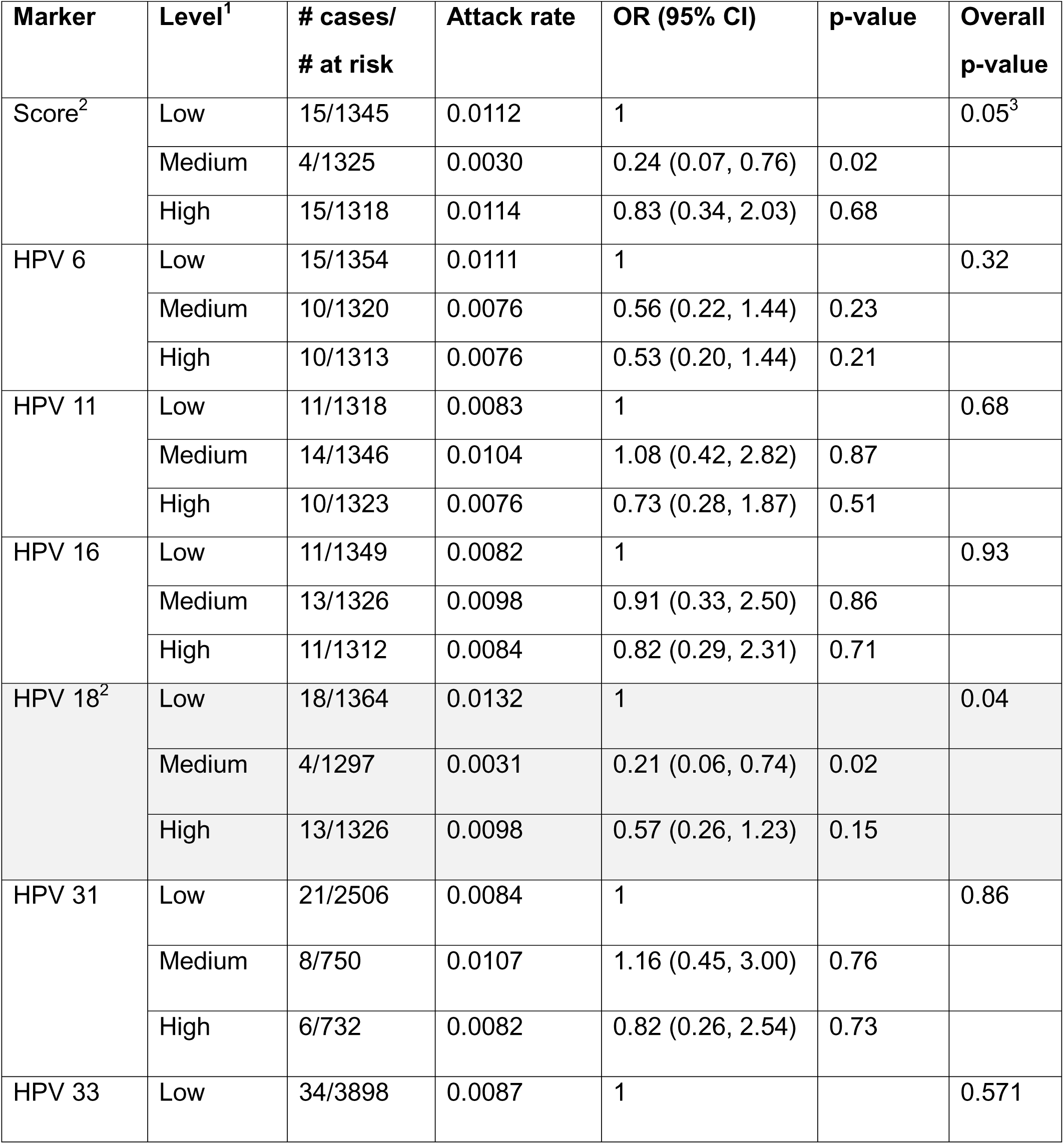
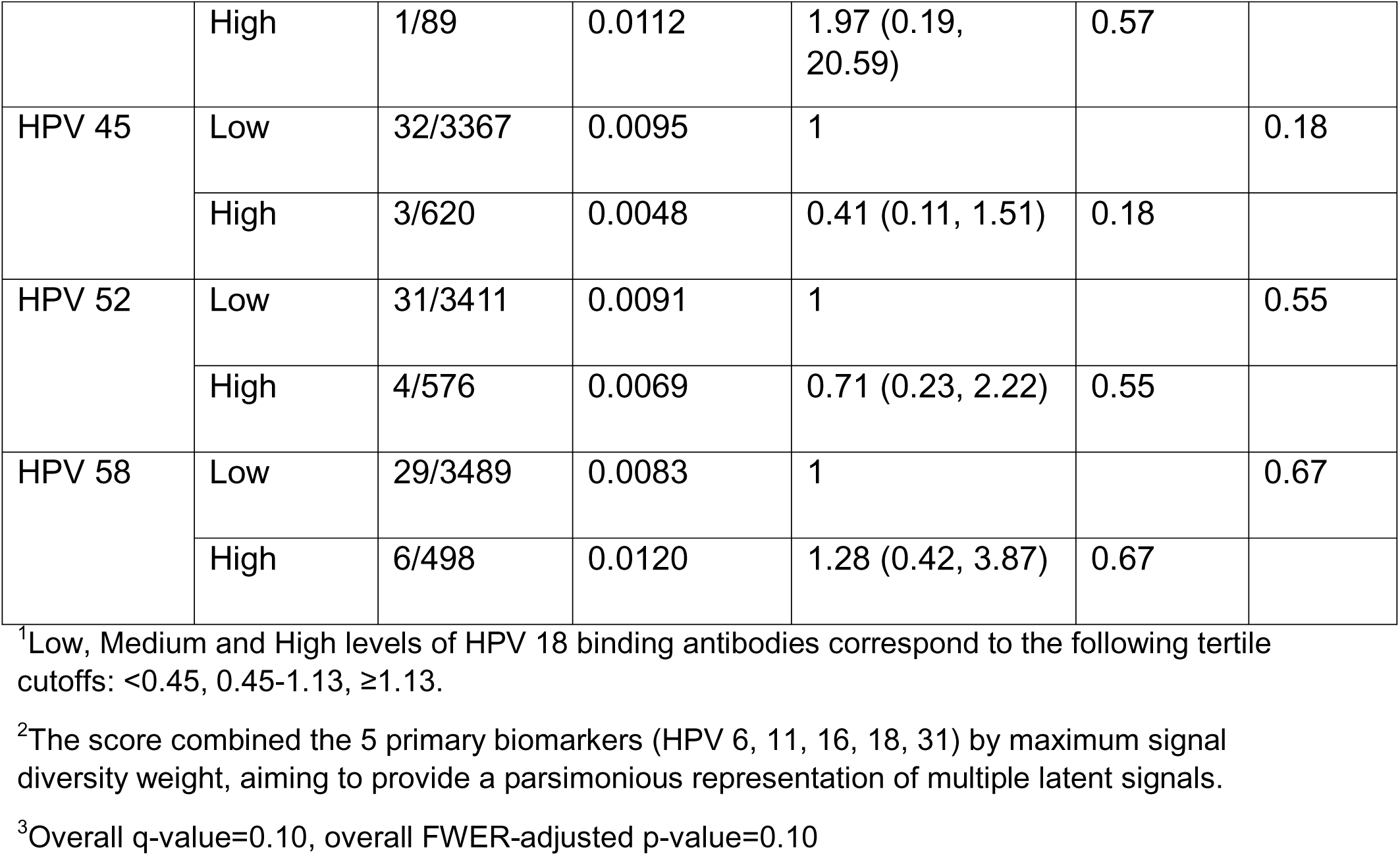
Categorical Month 18 IgG binding antibody markers as correlates of risk of Any HPV in the main analysis population (eligible and enrolled in cervical sampling cohort after Month 18). Odds ratios (ORs) are reported from univariate logistic regression models of the marker, adjusted for age > 14 and accounting for case-control sampling via inverse sampling probability weights. Shaded rows indicate overall p-value<0.05.

| Marker | Level<sup>1</sup> | # cases/<br># at risk | Attack rate | OR (95% CI) | p-value | Overall<br>p-value |
| --- | --- | --- | --- | --- | --- | --- |
| Score<sup>2</sup> | Low | 15/1345 | 0.0112 | 1 |  | 0.05<sup>3</sup> |
|  | Medium | 4/1325 | 0.0030 | 0.24 (0.07, 0.76) | 0.02 |  |
|  | High | 15/1318 | 0.0114 | 0.83 (0.34, 2.03) | 0.68 |  |
| HPV 6 | Low | 15/1354 | 0.0111 | 1 |  | 0.32 |
|  | Medium | 10/1320 | 0.0076 | 0.56 (0.22, 1.44) | 0.23 |  |
|  | High | 10/1313 | 0.0076 | 0.53 (0.20, 1.44) | 0.21 |  |
| HPV 11 | Low | 11/1318 | 0.0083 | 1 |  | 0.68 |
|  | Medium | 14/1346 | 0.0104 | 1.08 (0.42, 2.82) | 0.87 |  |
|  | High | 10/1323 | 0.0076 | 0.73 (0.28, 1.87) | 0.51 |  |
| HPV 16 | Low | 11/1349 | 0.0082 | 1 |  | 0.93 |
|  | Medium | 13/1326 | 0.0098 | 0.91 (0.33, 2.50) | 0.86 |  |
|  | High | 11/1312 | 0.0084 | 0.82 (0.29, 2.31) | 0.71 |  |
| HPV 18<sup>2</sup> | Low | 18/1364 | 0.0132 | 1 |  | 0.04 |
|  | Medium | 4/1297 | 0.0031 | 0.21 (0.06, 0.74) | 0.02 |  |
|  | High | 13/1326 | 0.0098 | 0.57 (0.26, 1.23) | 0.15 |  |
| HPV 31 | Low | 21/2506 | 0.0084 | 1 |  | 0.86 |
|  | Medium | 8/750 | 0.0107 | 1.16 (0.45, 3.00) | 0.76 |  |
|  | High | 6/732 | 0.0082 | 0.82 (0.26, 2.54) | 0.73 |  |
| HPV 33 | Low | 34/3898 | 0.0087 | 1 |  | 0.571 |
|  | High | 1/89 | 0.0112 | 1.97 (0.19, 20.59) | 0.57 |  |
| HPV 45 | Low | 32/3367 | 0.0095 | 1 |  | 0.18 |
|  | High | 3/620 | 0.0048 | 0.41 (0.11, 1.51) | 0.18 |  |
| HPV 52 | Low | 31/3411 | 0.0091 | 1 |  | 0.55 |
|  | High | 4/576 | 0.0069 | 0.71 (0.23, 2.22) | 0.55 |  |
| HPV 58 | Low | 29/3489 | 0.0083 | 1 |  | 0.67 |
|  | High | 6/498 | 0.0120 | 1.28 (0.42, 3.87) | 0.67 |  |

<sup>1</sup>Low, Medium and High levels of HPV 18 binding antibodies correspond to the following tertile cutoffs: <0.45, 0.45-1.13, ≥1.13.
<sup>2</sup>The score combined the 5 primary biomarkers (HPV 6, 11, 16, 18, 31) by maximum signal diversity weight, aiming to provide a parsimonious representation of multiple latent signals.
<sup>3</sup>Overall q-value=0.10, overall FWER-adjusted p-value=0.10

Neutralizing antibody titers showed no significant association with Any HPV in either analysis population for any HPV type considered (Tables S3-S6). There was a trend seen for HPV 52 titers in the alternative analysis population (continuous titer OR=3.84, 95%CI=(1.02, 14.44), p=0.047 for 10-fold increase in HPV 52 ID50 titer; OR=1.47, 95%CI=(0.49, 4.47), p=0.493 for medium vs. low levels of HPV 52 and OR=3.12, 95%CI=(1.23, 7.89), p=0.016 for high vs. low levels of HPV 52), Tables S5-S6. However, this was not seen in the main analysis population for either continuous or categorical HPV 52 neutralizing antibody titers (Tables S3-S4).

## Discussion

Higher month 18 HPV 18 binding antibody titers were associated with a lower risk of persistent HPV infection in the IARC India study. These results held for both continuous and categorical marker levels in the main analysis population (eligible and enrolled in the cervical sampling cohort after month 18) and in the alternative analysis population of participants susceptible after Month 18. Neutralizing antibody titers to HPV 18, however, were not associated with persistent HPV infection in either population nor were binding or neutralizing antibodies to any other HPV type considered.

Our study had several limitations, including the small number of vaccine-targeted persistent infections and the assumed absence of HPV exposure prior to cervical sampling following marriage or childbirth. Given its known cross-reactivity, HPV 31 was included in the primary outcome of any persistent HPV infection in addition to the 4 vaccine-targeted HPV types to increase the number of cases available for analysis [13–16]. However, vaccine-targeted antibodies might not be expected to be as a strong correlate against HPV 31 persistent infections as they would be against vaccine-targeted infections. To mitigate the second limitation of the possibility of HPV exposure at the time of blood sample collection at month 18, we repeated the correlates analysis in the cohort of participants who were married or pregnant after month 18. The results were very similar to those in the main analysis population.

Our descriptive analysis and the reported association between increased HPV 18 titers and reduced risk of any persistent HPV infection support the hypothesis that HPV 18 titers correlate with HPV 18 persistent infection. After HPV 31, type 18 was the most frequently observed persistent infection in our cohort. Once a greater number of HPV 18 persistent infections accrue, the type-specific correlates analysis can be conducted to assess this hypothesis.

Similarly, once more cases of any vaccine-targeted persistent infections accrue, the correlates analysis could be conducted on these alone to gain further insight into the role of vaccine-induced immune responses and protection from vaccine-targeted HPV. Our findings suggest this is highly plausible.

While a protective antibody titer threshold has not yet been identified for HPV, regulatory agencies, including the FDA, rely on immunogenicity as a key factor in granting vaccine licensure. The findings reported in this study represent the first evidence of a putative correlate of HPV risk, critical to the understanding, development and accelerated evaluation of new vaccine candidates.

## Data Availability

All data included in the present study will be made available upon reasonable request to the authors.

## Author Confirmation

All authors attest they meet the ICMJE criteria for authorship.

## Contributions

Zoe Moodie: Conceptualization, methodology, formal analysis, investigation, writing – original draft, visualization, supervision

Bhavesh Borate: formal analysis, investigation, visualization

Youyi Fong: methodology, formal analysis, investigation, visualization

Partha Basu: Conceptualization; Funding acquisition; Investigation; Methodology;

Resources; Supervision; Validation; Visualization; Writing—review & editing

Richard Muwonge: Data curation; Formal analysis; Writing—review & editing

## Acknowledgments

The authors thank Eric Lucas (IARC) for data support, Solmaz Shortobani and Drienna Holman (Fred Hutch) for project management, and Dr. Peter Dull for helpful discussion.

## Supplemental Material

**Figure S1:**
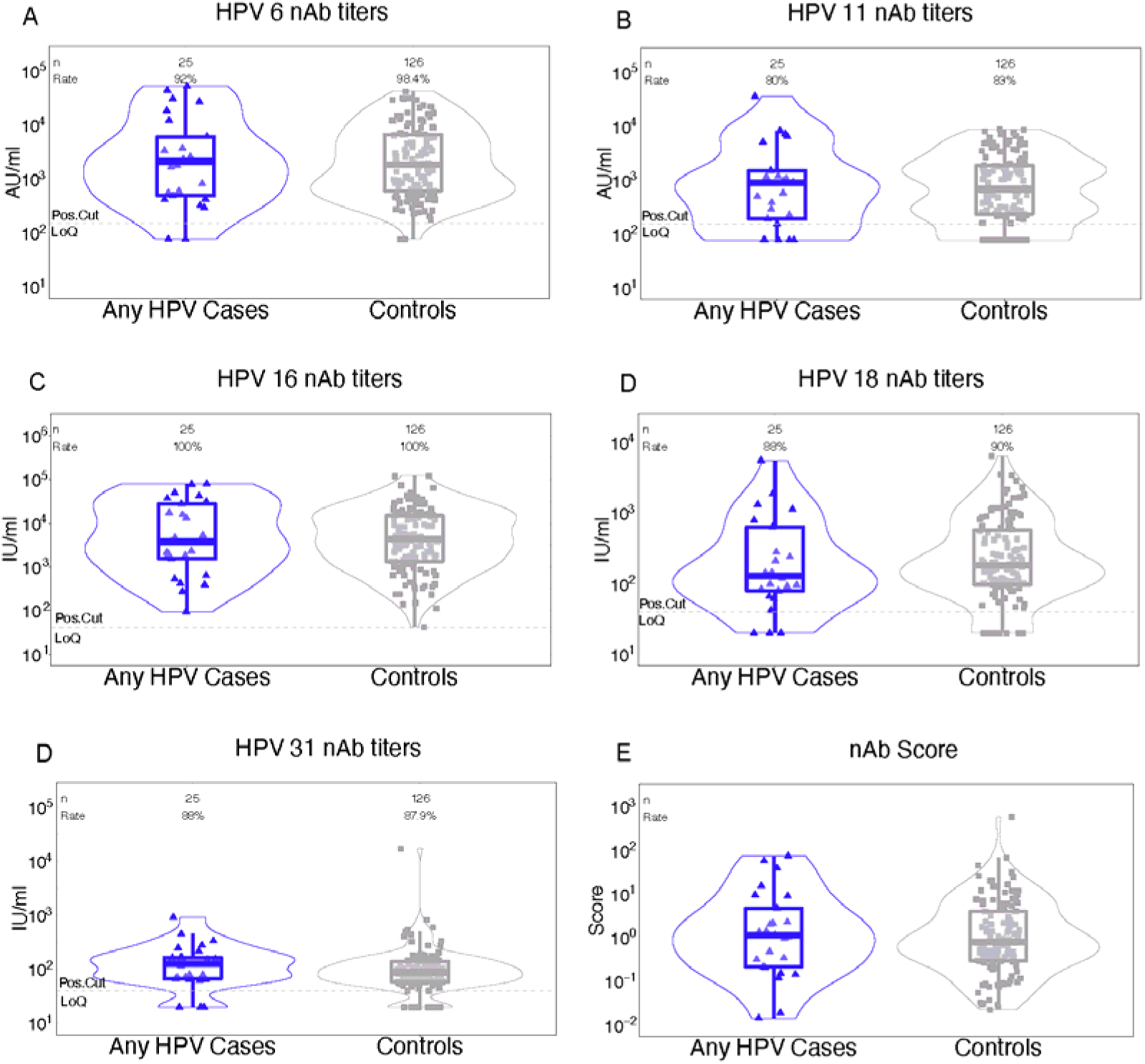
Distribution of Month 18 neutralizing antibody titers to HPV 6, 11, 16, 18, 31 and the score for Any HPV cases and controls in the main analysis population. The number of participants is denoted by n; Rate refers to the proportion of participants with a positive response; Pos. Cut denotes the positivity cutoff; LoQ denotes the limit of quantitation.

**Figure S2:**
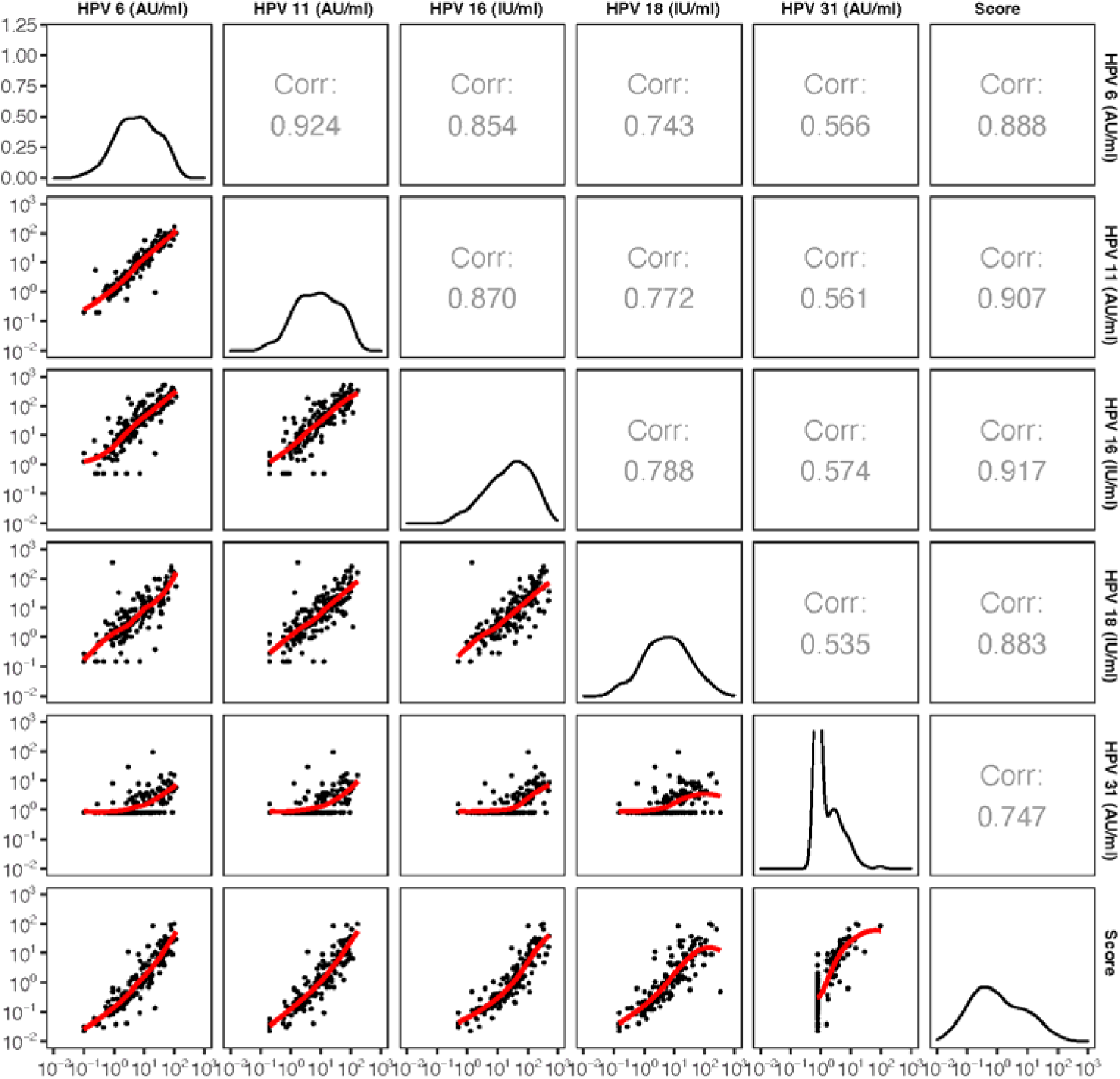
Intercorrelation among Month 18 IgG binding antibody titers to HPV 6, 11, 16, 18, 31, and score. Weighted Spearman pairwise correlations are noted in the upper diagonal tiles, smoothed binding antibody distributions are shown along the diagonal, and scatterplots are shown in the lower diagonal tiles with red lines representing the Lowess smoother.

**Figure S3:**
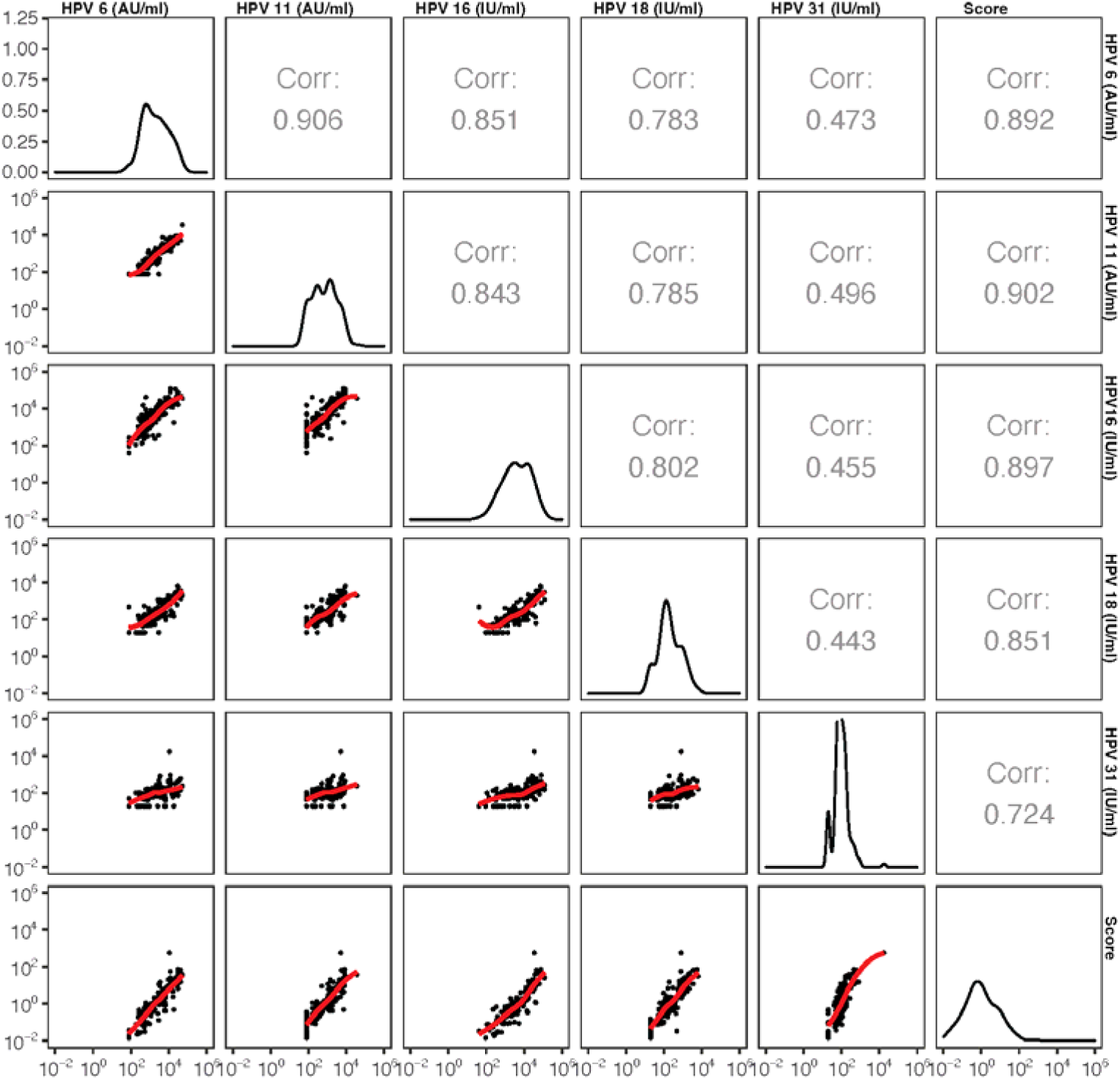
Intercorrelation among Month 18 neutralizing antibody titers to HPV 6, 11, 16, 18, and score. Weighted Spearman pairwise correlations are noted in the upper diagonal tiles, smoothed binding antibody distributions are shown along the diagonal, and scatterplots are shown in the lower diagonal tiles with red lines representing the Lowess smoother.

**Figure S4:**
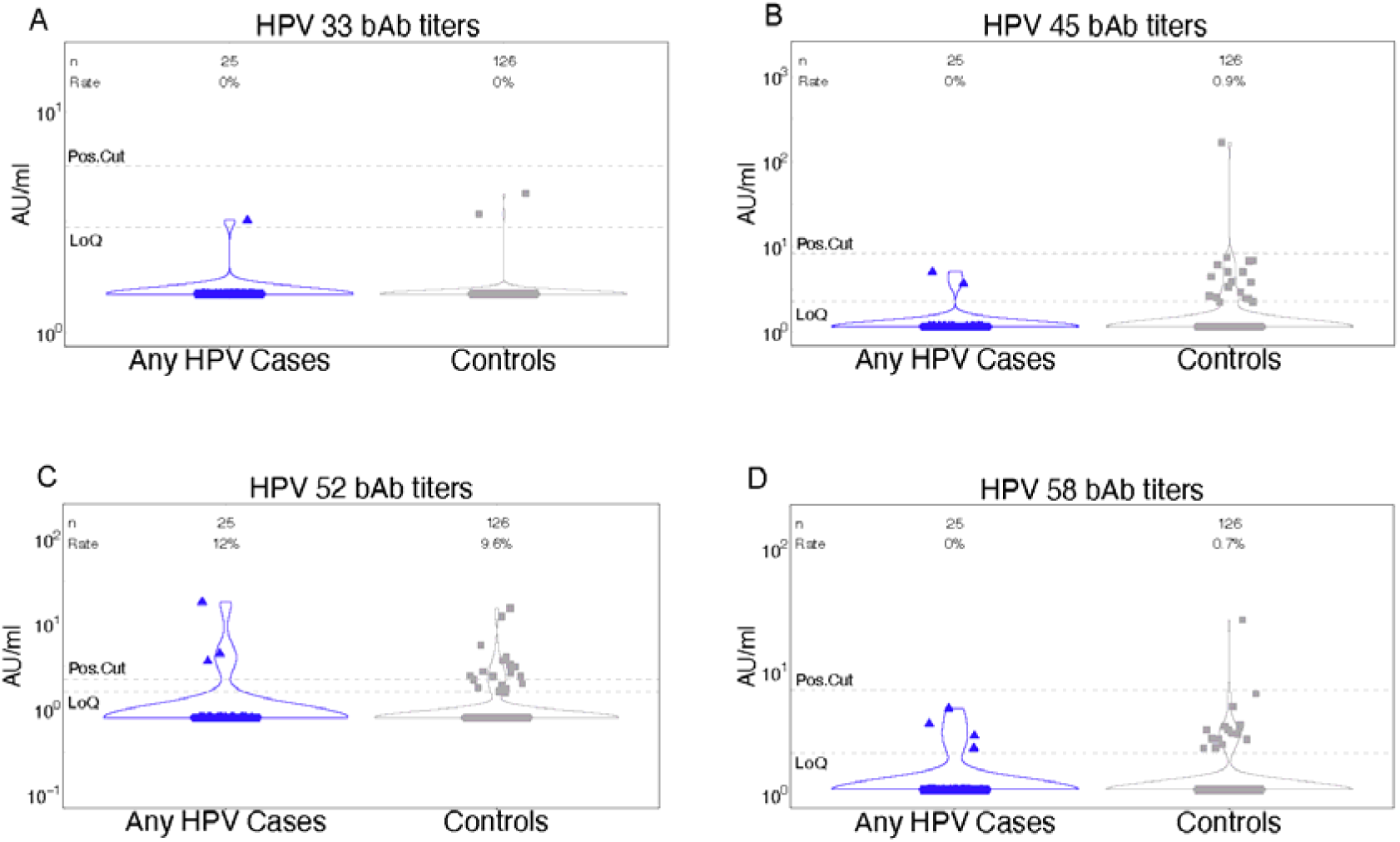
Distribution of Month 18 binding antibody titers to HPV 33, 45, 52, 58 for Any HPV cases and controls in main analysis population. The number of participants is denoted by n; Rate refers to the proportion of participants with a positive response; Pos. Cut denotes the positivity cutoff; LoQ denotes the limit of quantitation.

**Figure S5:**
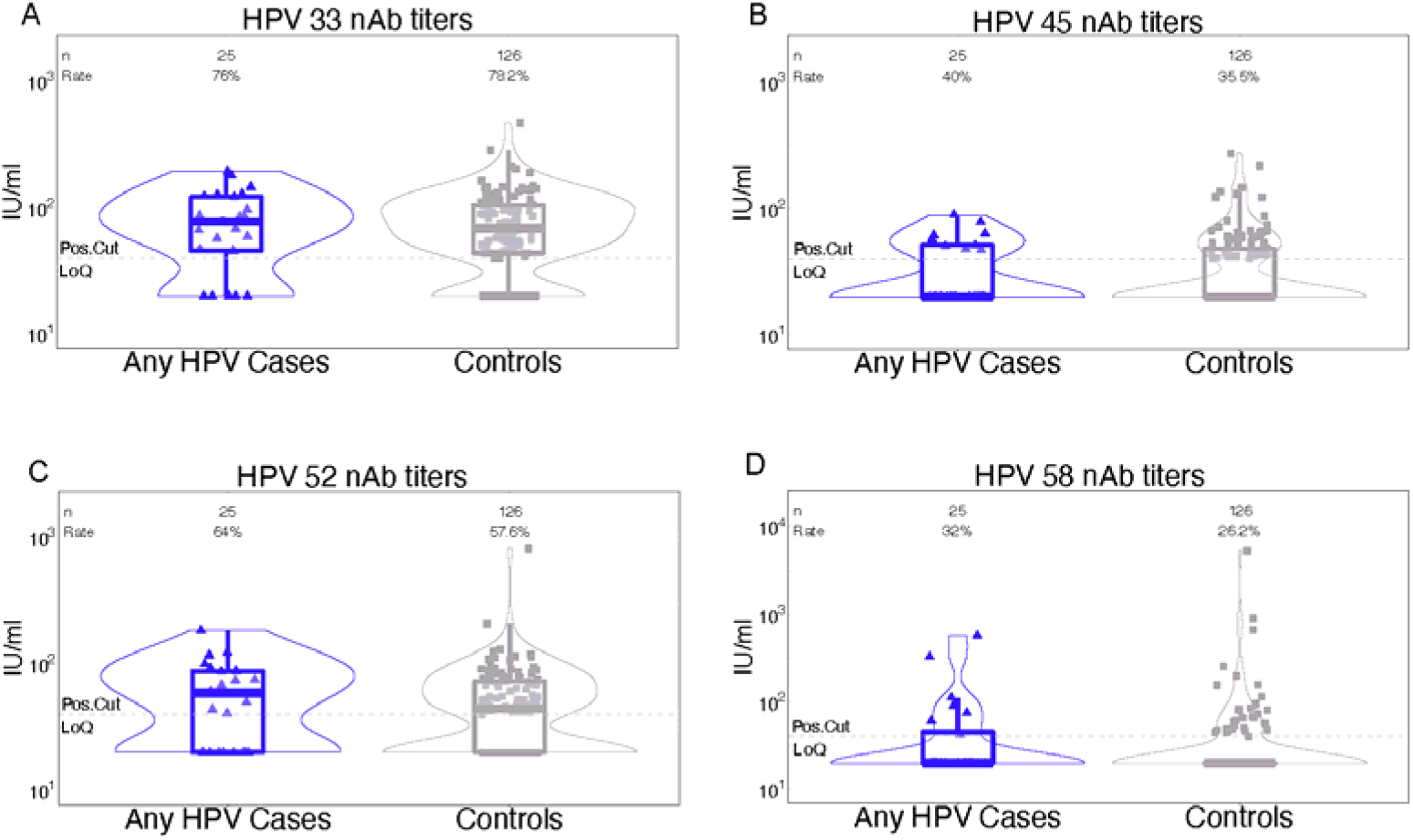
Distribution of Month 18 neutralizing antibody titers to HPV 33, 45, 52, 58 for Any HPV cases and controls in main analysis population. The number of participants is denoted by n; Rate refers to the proportion of participants with a positive response; Pos. Cut denotes the positivity cutoff; LoQ denotes the limit of quantitation.

**Table S1:** Baseline demographics and case-control status of per-protocol cohort in main analysis population.

|  | **Single dose**<br>**(n=1248)** | **Two doses**<br>**(n=1904)** | **Three doses**<br>**(n=858)** | **Total**<br>**(n=4010)** |
| --- | --- | --- | --- | --- |
| **Age (in years)** |  |  |  |  |
| **<=14** | 504 | 766 | 369 | 1639 |
| **>14** | 744 | 1138 | 489 | 2371 |
| **Mean (range)** | 15 (10-18) | 15 (10-18) | 15 (10-18) | 15 (10-18) |
| **Site** |  |  |  |  |
| **Ahmedabad** | 166 | 0 | 0 | 166 |
| **Ambilikkai** | 46 | 173 | 111 | 330 |
| **Barshi** | 851 | 1100 | 240 | 2191 |
| **Delhi** | 7 | 80 | 51 | 138 |
| **Hyderabad** | 48 | 47 | 0 | 95 |
| **Mizoram** | 2 | 17 | 14 | 33 |
| **Mumbai** | 2 | 20 | 0 | 22 |
| **Pune** | 125 | 462 | 436 | 1023 |
| **Sikkim** | 1 | 5 | 6 | 12 |
| **Cases** |  |  |  |  |
| **Any HPV** | 13 | 15 | 7 | 35 |
| **HPV 6** | 1 | 2 | 1 | 4 |
| **HPV 11** | 0 | 1 | 1 | 2 |
| **HPV 16** | 1 | 1 | 0 | 2 |
| **HPV 18** | 2 | 5 | 2 | 9 |
| **HPV 31** | 9 | 6 | 3 | 18 |
| **Controls** | 1235 | 1889 | 851 | 3975 |

**Table S2:** Continuous IgG binding antibody markers as correlates of risk of Any HPV in the alternative analysis population (susceptible after Month 18). Odds ratios (ORs) are reported from univariate logistic regression models of the marker, adjusted for age > 14 and accounting for case-control sampling via inverse sampling probability weights. Shaded row indicates p-value<0.05 (p-value=0.047 prior to rounding).

| Marker | # cases/<br># at risk | OR per 10-fold increment<br>(95% CI) | p-value |
| --- | --- | --- | --- |
| Score<sup>1</sup> | 26/3201 | 0.77 (0.47, 1.26) | 0.29<sup>2</sup> |
| HPV 6 | 26/3201 | 0.68 (0.38, 1.22) | 0.20 |
| HPV 11 | 26/3201 | 0.86 (0.49, 1.50) | 0.60 |
| HPV 16 | 26/3201 | 0.76 (0.43, 1.34) | 0.34 |
| HPV 18 | 26/3201 | 0.58 (0.34, 0.99) | 0.05 |
| HPV 31 | 26/3201 | 0.89 (0.33, 2.41) | 0.83 |
| HPV 33 | 26/3201 | 4.01 (0.01, >100) | 0.66 |
| HPV 45 | 26/3201 | 0.26 (0.03, 2.22) | 0.22 |
| HPV 52 | 26/3201 | 0.46 (0.07, 3.11) | 0.43 |
| HPV 58 | 26/3201 | 2.41 (0.25, 23.00) | 0.45 |

<sup>1</sup>The score combined the 5 primary biomarkers (HPV 6, 11, 16, 18, 31) by maximum signal diversity weight, aiming to provide a parsimonious representation of multiple latent signals.
<sup>2</sup>Overall q-value=0.29, overall FWER-adjusted p-value=0.29

**Table S3:** Categorical IgG binding antibody markers as correlates of risk of Any HPV in the alternative analysis population (susceptible after Month 18). Odds ratios (ORs) are reported from univariate logistic regression models of the marker, adjusted for age > 14 and accounting for case-control sampling via inverse sampling probability weights. Shaded rows indicate marker with p-value<0.05 for continuous IgG binding antibody marker (Table S1).

| Marker | Level<sup>1</sup> | # cases/<br># at risk | Attack rate | OR (95% CI) | p-value | Overall<br>p-value |
| --- | --- | --- | --- | --- | --- | --- |
| Score<sup>2</sup> | Low | 13/1,083 | 0.0120 | 1 |  | 0.10<sup>3</sup> |
|  | Medium | 4/1,061 | 0.0038 | 0.26 (0.07, 0.90) | 0.03 |  |
|  | High | 9/1,057 | 0.0085 | 0.69 (0.26, 1.80) | 0.44 |  |
| HPV 6 | Low | 13/1,052 | 0.0124 | 1 |  | 0.20 |
|  | Medium | 7/1,083 | 0.0065 | 0.52 (0.20, 1.38) | 0.19 |  |
|  | High | 6/1,066 | 0.0056 | 0.43 (0.14, 1.27) | 0.13 |  |
| HPV 11 | Low | 9/1,072 | 0.0084 | 1 |  | 0.79 |
|  | Medium | 9/1,080 | 0.0083 | 0.96 (0.34, 2.71) | 0.94 |  |
|  | High | 7/1,049 | 0.0067 | 0.72 (0.26, 1.96) | 0.52 |  |
| HPV 16 | Low | 13/1,068 | 0.0122 | 1 |  | 0.17 |
|  | Medium | 6/1,104 | 0.0054 | 0.38 (0.14, 1.07) | 0.07 |  |
|  | High | 7/1,029 | 0.0068 | 0.51 (0.18, 1.44) | 0.20 |  |
| HPV 18<sup>2</sup> | Low | 14/1,100 | 0.0134 | 1 |  | 0.06 |
|  | Medium | 2/1,058 | 0.0023 | 0.17 (0.02-0.65) | 0.004 |  |
|  | High | 9/1,043 | 0.0092 | 0.64 (0.27, 1.55) | 0.33 |  |
| HPV 31 | Low | 17/1,956 | 0.0087 | 1 |  | 0.37 |
|  | Medium | 7/629 | 0.0111 | 1.27 (0.49, 3.26) | 0.62 |  |
|  | High | 2/615 | 0.0033 | 0.43 (0.12, 1.59) | 0.21 |  |
| HPV 33 | Low | 25/3,131 | 0.0080 | 1 |  | 0.53 |
|  | High | 1/70 | 0.0143 | 2.16 (0.20, 23.39) | 0.53 |  |
| HPV 45 | Low | 24/2,613 | 0.0092 | 1 |  | 0.19 |
|  | High | 2/588 | 0.0034 | 0.40 (0.11, 1.55) | 0.19 |  |
| HPV 52 | Low | 24/2,676 | 0.0090 | 1 |  | 0.25 |
|  | High | 2/525 | 0.0038 | 0.48 (0.13, 1.70) | 0.25 |  |
| HPV 58 | Low | 21/2,806 | 0.0075 | 1 |  | 0.44 |
|  | High | 5/395 | 0.0127 | 1.60 (0.49, 5.21) | 0.44 |  |

<sup>1</sup>Low, Medium and High levels of HPV 18 binding antibodies correspond to the following tertile cutoffs: <0.45, 0.45-1.12, ≥1.12.
<sup>2</sup>The score combined the 5 primary biomarkers (HPV 6, 11, 16, 18, 31) by maximum signal diversity weight, aiming to provide a parsimonious representation of multiple latent signals.
<sup>3</sup>Overall q-value=0.21, overall FWER-adjusted p-value=0.21

**Table S4:** Continuous IgG neutralizing antibody markers as correlates of risk of Any HPV in the main analysis population (eligible and enrolled in cervical sampling cohort after Month 18). Odds ratios (ORs) are reported from univariate logistic regression models of the marker, adjusted for age > 14 and accounting for case-control sampling via inverse sampling probability weights.

| **Marker** | **# cases/**<br>**# at risk** | **OR per 10-fold increment**<br>**(95% CI)** | **p-value** |
| --- | --- | --- | --- |
| Score | 35/3,987 | 1.00 (0.64, 1.56) | 1.00<sup>1</sup> |
| HPV 6 | 35/3,987 | 0.90 (0.47, 1.70) | 0.74 |
| HPV 11 | 35/3,987 | 0.99 (0.53, 1.88) | 0.99 |
| HPV 16 | 35/3,987 | 0.98 (0.55, 1.73) | 0.94 |
| HPV 18 | 35/3,987 | 0.68 (0.36, 1.28) | 0.23 |
| HPV 31 | 35/3,987 | 1.49 (0.63, 3.51) | 0.36 |
| HPV 33 | 35/3,987 | 0.85 (0.28, 2.57) | 0.77 |
| HPV 45 | 35/3,987 | 0.88 (0.27, 2.88) | 0.83 |
| HPV 52 | 35/3,987 | 2.09 (0.68, 6.43) | 0.20 |
| HPV 58 | 35/3,987 | 1.43 (0.66, 3.10) | 0.36 |

<sup>1</sup>Overall q-value=1.00, overall FWER-adjusted p-value=1.00

**Table S5:**
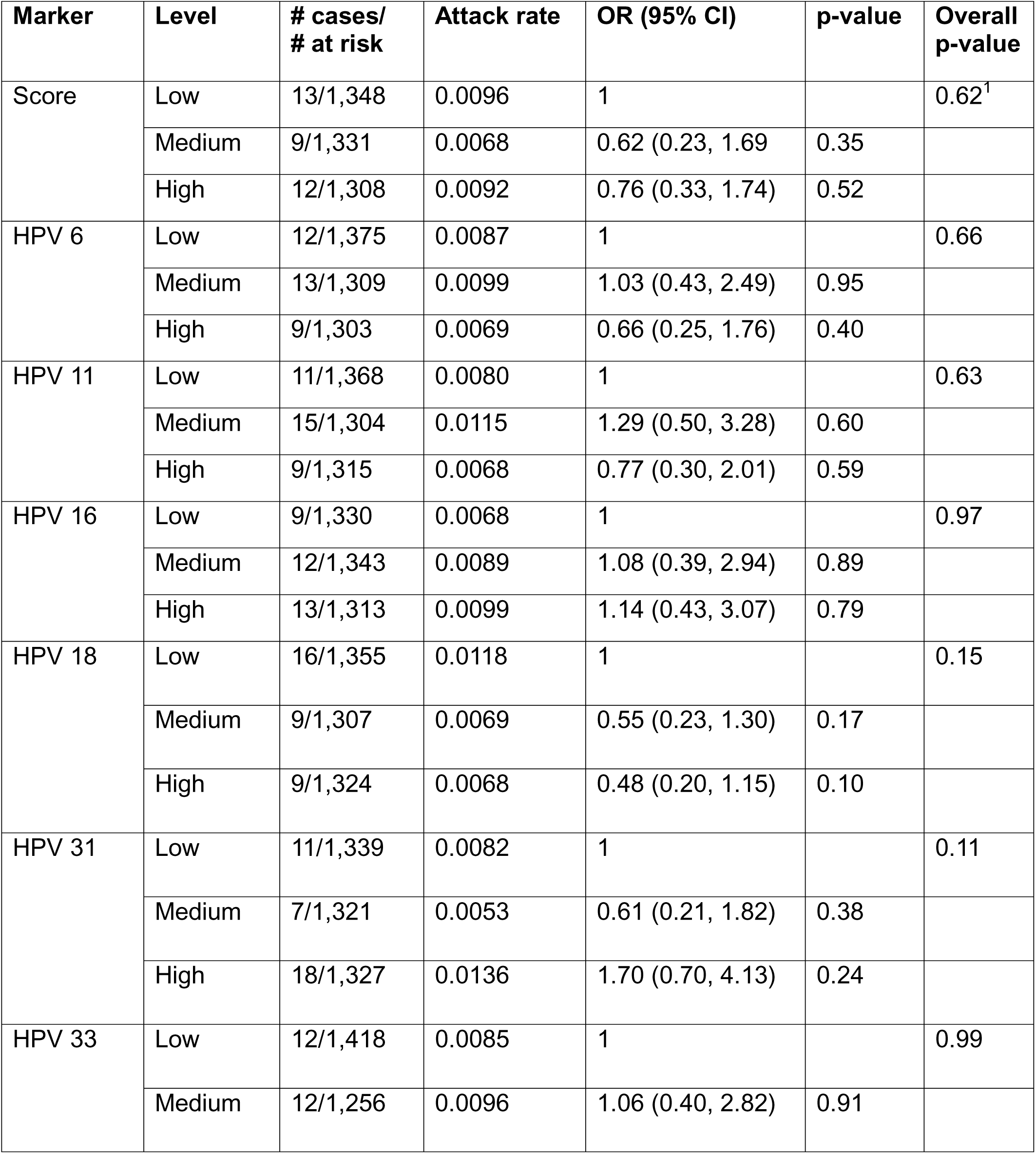
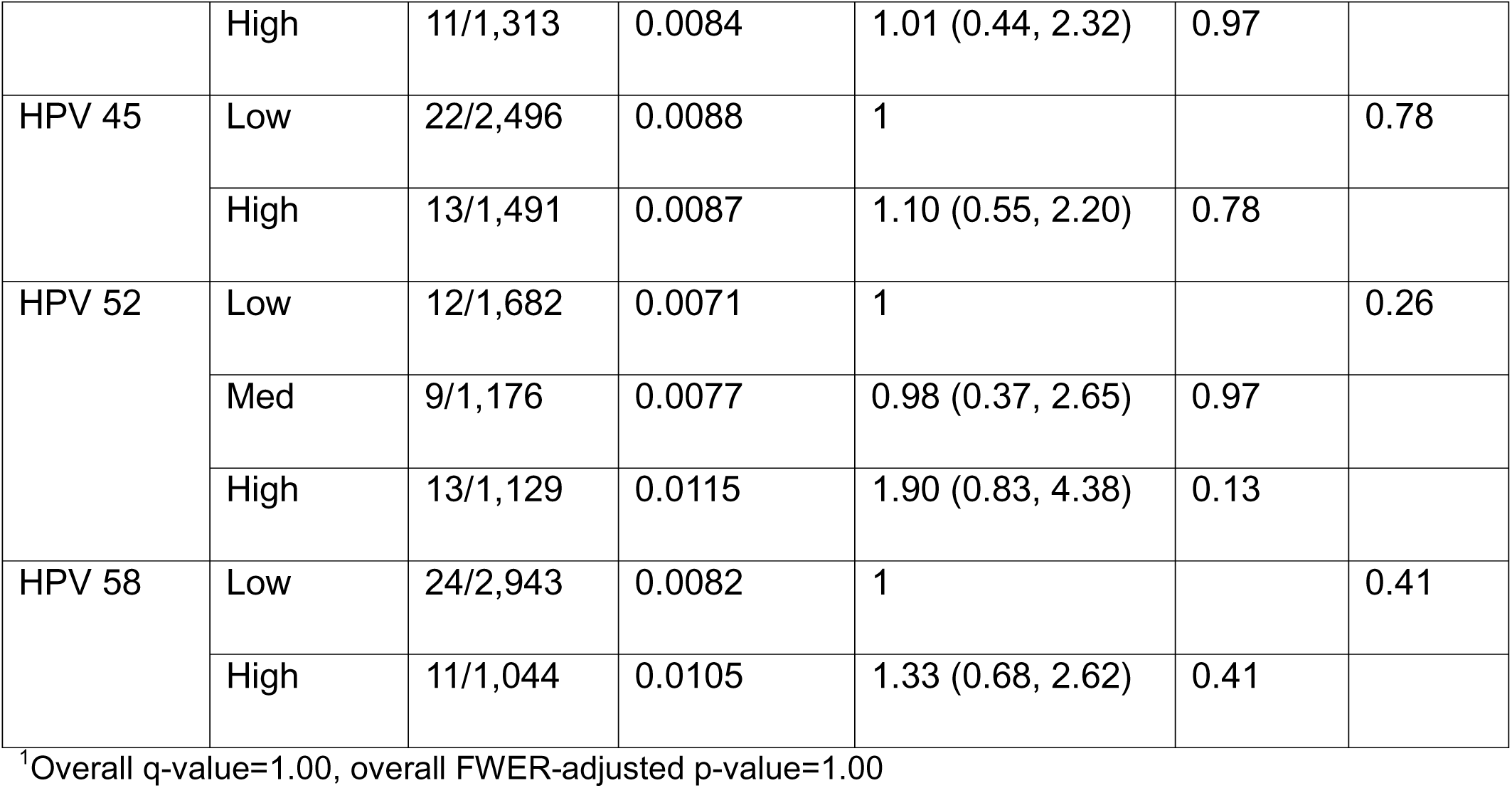
Categorical IgG neutralizing antibody markers as correlates of risk of Any HPV in the main analysis population (eligible and enrolled in cervical sampling cohort after Month 18). Odds ratios (ORs) are reported from univariate logistic regression models of the marker, adjusted for age > 14 and accounting for case-control sampling via inverse sampling probability weights.

**Table S6:** Continuous IgG neutralizing antibody markers as correlates of risk of Any HPV in the alternative analysis population (susceptible after Month 18). Odds ratios (ORs) are reported from univariate logistic regression models of the marker, adjusted for age > 14 and accounting for case-control sampling via inverse sampling probability weights. Shaded row indicates p-value<0.05 (p-value=0.047 prior to rounding).

| Marker | # cases/<br># at risk | OR per 10-fold increment<br>(95% CI) | p-value |
| --- | --- | --- | --- |
| Score | 26/3,201 | 1.06 (0.64, 1.77) | 0.81<sup>1</sup> |
| HPV 6 | 26/3,201 | 0.92 (0.46, 1.84) | 0.81 |
| HPV 11 | 26/3,201 | 1.10 (0.55, 2.20) | 0.79 |
| HPV 16 | 26/3,201 | 0.94 (0.49, 1.79) | 0.84 |
| HPV 18 | 26/3,201 | 0.76 (0.35, 1.62) | 0.47 |
| HPV 31 | 26/3,201 | 1.74 (0.71, 4.27) | 0.22 |
| HPV 33 | 26/3,201 | 1.85 (0.53, 6.45) | 0.34 |
| HPV 45 | 26/3,201 | 1.41 (0.40, 4.97) | 0.59 |
| HPV 52 | 26/3,201 | 3.84 (1.02, 14.44) | 0.05 |
| HPV 58 | 26/3,201 | 2.54 (0.86, 7.53) | 0.09 |

<sup>1</sup>Overall q-value=0.81, overall FWER-adjusted p-value=1.00

**Table S7:** Categorical IgG neutralizing antibody markers as correlates of risk of Any HPV in the alternative analysis population (susceptible after Month 18). Odds ratios (ORs) are reported from univariate logistic regression models of the marker, adjusted for age > 14 and accounting for case-control sampling via inverse sampling probability weights.

| Marker | Level | # cases/<br># at risk | Attack rate | OR (95% CI) | p-value | Overall<br>p-value |
| --- | --- | --- | --- | --- | --- | --- |
| Score | Low | 11/1,085 | 0.0101 | 1 |  | 0.511<sup>1</sup> |
|  | Medium | 6/1,065 | 0.0056 | 0.55 (0.20, 1.52) | 0.25 |  |
|  | High | 9/1,051 | 0.0086 | 0.88 (0.37, 2.11) | 0.78 |  |
| HPV 6 | Low | 9/1,095 | 0.0082 | 1 |  | 0.80 |
|  | Medium | 9/1,054 | 0.0085 | 1.07 (0.40, 2.89) | 0.89 |  |
|  | High | 7/1,052 | 0.0067 | 0.74 (0.26, 2.07) | 0.57 |  |
| HPV 11 | Low | 8/1,134 | 0.0071 | 1 |  | 0.72 |
|  | Medium | 11/1,025 | 0.0107 | 1.44 (0.50, 4.15) | 0.50 |  |
|  | High | 7/1,042 | 0.0067 | 0.93 (0.33, 2.57) | 0.88 |  |
| HPV 16 | Low | 9/1,069 | 0.0084 | 1 |  | 0.76 |
|  | Medium | 7/1,089 | 0.0064 | 0.68 (0.23, 2.02) | 0.49 |  |
|  | High | 9/1,043 | 0.0086 | 0.94 (0.34, 2.63) | 0.91 |  |
| HPV 18<sup>2</sup> | Low | 12/1,070 | 0.0112 | 1 |  | 0.33 |
|  | Medium | 7/1,073 | 0.0065 | 0.60 (0.24, 1.50) | 0.28 |  |
|  | High | 7/1,059 | 0.0066 | 0.55 (0.21, 1.45) | 0.23 |  |
| HPV 31 | Low | 7/1,074 | 0.0065 | 1 |  | 0.10 |
|  | Medium | 6/1,093 | 0.0055 | 0.81 (0.26, 2.46) | 0.71 |  |
|  | High | 13/1,034 | 0.0126 | 2.03 (0.77, 5.32) | 0.15 |  |
| HPV 33 | Low | 6/1,090 | 0.0055 | 1 |  | 0.35 |
|  | Medium | 12/977 | 0.0123 | 2.13 (0.72, 6.32) | 0.18 |  |
|  | High | 8/1,134 | 0.0071 | 1.41 (0.49, 4.08) | 0.52 |  |
| HPV 45 | Low | 14/2,012 | 0.0070 | 1 |  | 0.28 |
|  | High | 12/1,189 | 0.0101 | 1.49 (0.72, 3.08) | 0.28 |  |
| HPV 52* | Low | 7/1,429 | 0.0049 | 1 |  | 0.053 |
|  | Medium | 7/886 | 0.0079 | 1.47 (0.49, 4.47) | 0.49 |  |
|  | High | 12/886 | 0.0135 | 3.12 (1.23, 7.89) | 0.02 |  |
| HPV 58 | Low | 17/2,393 | 0.0071 | 1 |  | 0.13 |
|  | High | 9/808 | 0.0111 | 1.78 (0.85, 3.75) | 0.13 |  |

<sup>1</sup>Overall q-value=0.81, overall FWER-adjusted p-value=1.00
<sup>2</sup>Low, Medium and High levels of HPV 52 binding antibodies correspond to the following tertile cutoffs: <1.3, 1.3-1.85, ≥1.85.

